# Convolutional Encoder-Decoder Networks for Volumetric Computed Tomography Surviews from Single- and Dual-View Topograms

**DOI:** 10.1101/2022.05.17.22275229

**Authors:** Nadav Shapira, Siddharth Bharthulwar, Peter B. Noël

## Abstract

Computed tomography (CT) is an extensively used imaging modality capable of generating detailed images of a patient’s internal anatomy for diagnostic and interventional procedures. High-resolution volumes are created by measuring and combining information along many radiographic projection angles. In current medical practice, single and dual-view two-dimensional (2D) topograms are utilized for planning the proceeding diagnostic scans and for selecting favorable acquisition parameters, either manually or automatically, as well as for dose modulation calculations. In this study, we develop modified 2D to three-dimensional (3D) encoder-decoder neural network architectures to generate CT-like volumes from single and dual-view topograms. We validate the developed neural networks on synthesized topograms from publicly available thoracic CT datasets. Finally, we assess the viability of the proposed transformational encoder-decoder architecture on both common image similarity metrics and quantitative clinical use case metrics, a first for 2D-to-3D CT reconstruction research. According to our findings, both single-input and dual-input neural networks are able to provide accurate volumetric anatomical estimates. The proposed technology will allow for improved (i) planning of diagnostic CT acquisitions, (ii) input for various dose modulation techniques, and (iii) recommendations for acquisition parameters and/or automatic parameter selection. It may also provide for an accurate attenuation correction map for positron emission tomography (PET) with only a small fraction of the radiation dose utilized.

## I. Introduction

X-ray computed tomography (CT) is an extensively used imaging modality that captures three-dimensional (3D) anatomical structures, primarily for diagnostic applications^1^. Clinical CT systems rely on measuring individual x-ray projections in axial or helical acquisition modes while rotating around the patient body, eliminating spatial disadvantages of prior two-dimensional (2D) radiographic x-ray modalities^2,3^. Through filtered back-projection (FBP) or state-of-the-art reconstruction techniques, such as iterative reconstruction (IR) or artificial intelligence (AI), modern CT scanners can produce high-resolution volumetric images of a patient’s anatomy^4^. However, by measuring hundreds of projections at varying angles, CT scans involve ionizing radiation exposure to patients, raising numerous health concerns such as damage to DNA and the associated increased risk of cancer^5–8^. Dose modulation techniques^9–11^ and manual or automatic kVp selection^12,13^ are examples of modern CT imaging methods that provide means to significantly reduce the amount of radiation required to generate datasets of diagnostic image quality. Dose modulation allows a reduction of radiation dose levels by adjusting the x-ray flux during a scan through current modulations of the accelerated electrons that bombard the anode. Such current modulations are based on patient-specific information that is derived from either coronal or sagittal 2D topograms, or both. Such 2D topograms, referred to by different CT vendors as scouts, surviews, or tomograms, provide input for both planning of the diagnostic scan and for estimating water-equivalent diameters (WED) that represent the patient size in every cross-section and serve as a basis for actively reducing/increasing the radiation dose in large/small cross sections^14^. Planning topograms are also being used to select the accelerating electron voltage (kVp), which can be determined either manually or automatically, in order to control the quality of the beam and ensure that a large proration of x-ray photons is utilized for imaging, e.g., higher kVp values for larger patient sizes^12,13,15^. Information in 3D-surview volumes that were generated from single- or dual-view topograms has the potential to improve current and future dose modulation techniques and acquisition parameter selections for reduced patient risk. For example, dynamic bowtie filter concepts for angle-dependent x-ray flux modulation^15–17^ would also greatly benefit from the additional information contained within 3D-surviews. Finally, 3D-surviews could enable improved planning of proceeding diagnostic scans, e.g., through better organ visualization.

In recent years the appeal of deep learning and AI in the medical imaging domain has inspired several studies investigating deep-learning facilitated CT reconstruction^18–20^. Notable studies involve transforming and denoising low-dose CT scans with convolutional autoencoders and convolutional neural network (CNN) facilitated limited-angle reconstruction^21–23^. Within the specific subfield of 2D-to-3D CT mapping, Shen *et al*. investigate deep learning methods for generating volumetric datasets from single-view radiographic projections^22^. However, few studies investigate the advantages of utilizing stereo 2D-to-3D CT. Katsen *et al*. developed a CNN to generate 3D knee bone segmentations from pairs of 2D knee radiographs^23^. However, the challenge of generating volumetric datasets for additional body sections, e.g., for thoracic CT scans, from more than one clinically obtainable projection remains.

In this study, we develop and implement single-view and dual-view CNNs for volumetric CT mapping from 2D topogram projections. Both neural networks follow a modified encoder-decoder architecture, as shown in Figure 1, and are comprised of three individual convolutional subnetworks: a feature representation subnetwork, a latent transformation subnetwork, and a generation subnetwork. To determine the theoretical viability of these encoder-decoder architectures, we train and assess their performance on publicly available CT datasets from the Lung Image Data Consortium dataset ^24^. Finally, we assess the clinical viability of the two neural networks with use case-specific metrics for common CT applications, including dose modulation techniques and improved clinical scan planning that is strengthened by superior anatomy-awareness.

**Figure 1:**
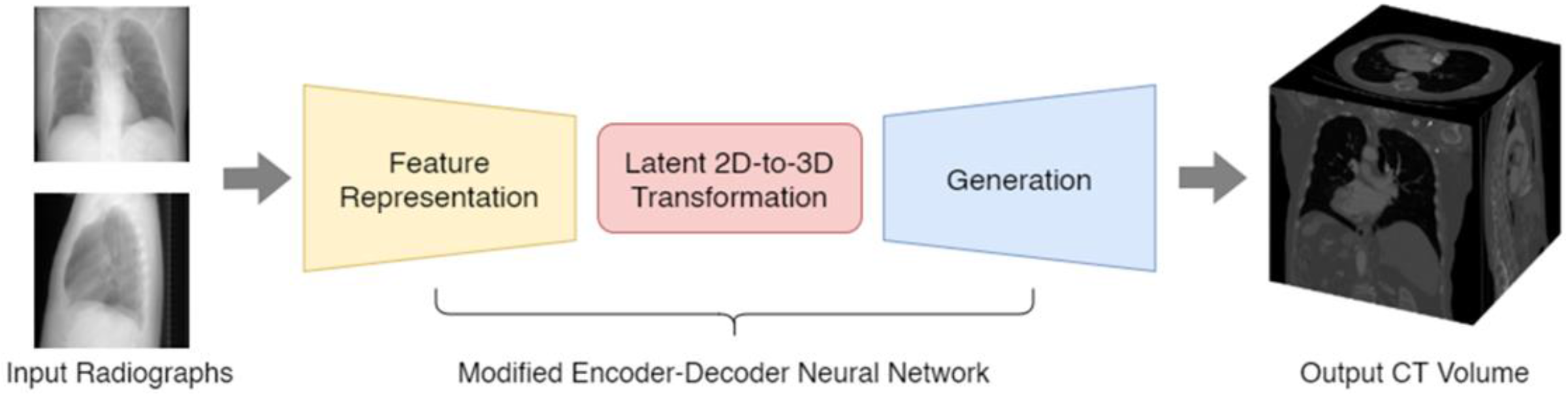
Schematic workflow diagram of the implemented encoder-decoder neural network architecture for single- or dual-view topogram (2D) to volume (3D) mapping. The workflow uses single- or dual-view topograms as input and includes a representation subnetworks (yellow), a transformation subnetwork (red), and a generation subnetwork (blue) to estimate CT-like volumetric outputs.

## II. Methods

### A. Data Collection and Generation of Paired Topograms

The publicly available Lung Image Data Consortium (LIDC) dataset includes images from 1050 helical thoracic CT examinations that is compiled from seven academic centers and eight different CT vendors^24^. Each exam contains both volumetric pixel information as well as relevant scan parameters (e.g., tube current and tube voltage). However, the database includes topograms for only a very limited number of cases, and typically only for a single view, i.e., sagittal or coronal. To accurately train deep neural networks that generate volumetric images from input 2D topograms, a large training dataset of paired 2D-3D data is required. For this, we implemented a dedicated synthetic topogram creation process^25^. The process, which is depicted in Figure 2, involves simulating multiple beams that travers through clinical CT volumes from a fixed x-ray point source. For each ray starting at the predefined point source location and entrance voxel, the next point of intersection is defined as the closest voxel border to the path of the ray. The process is repeated with the point of intersection representing the new entrance point, until the ray exits the volume. For each ray, the products of distance between entrance and exit point and the voxel attenuation are aggregated. Topogram intensities are then synthesized on a simulated 1024 × 1024 pixel x-ray detector by utilizing the Beer-Lambert Law of x-ray attenuation^5^:

**Figure 2:**
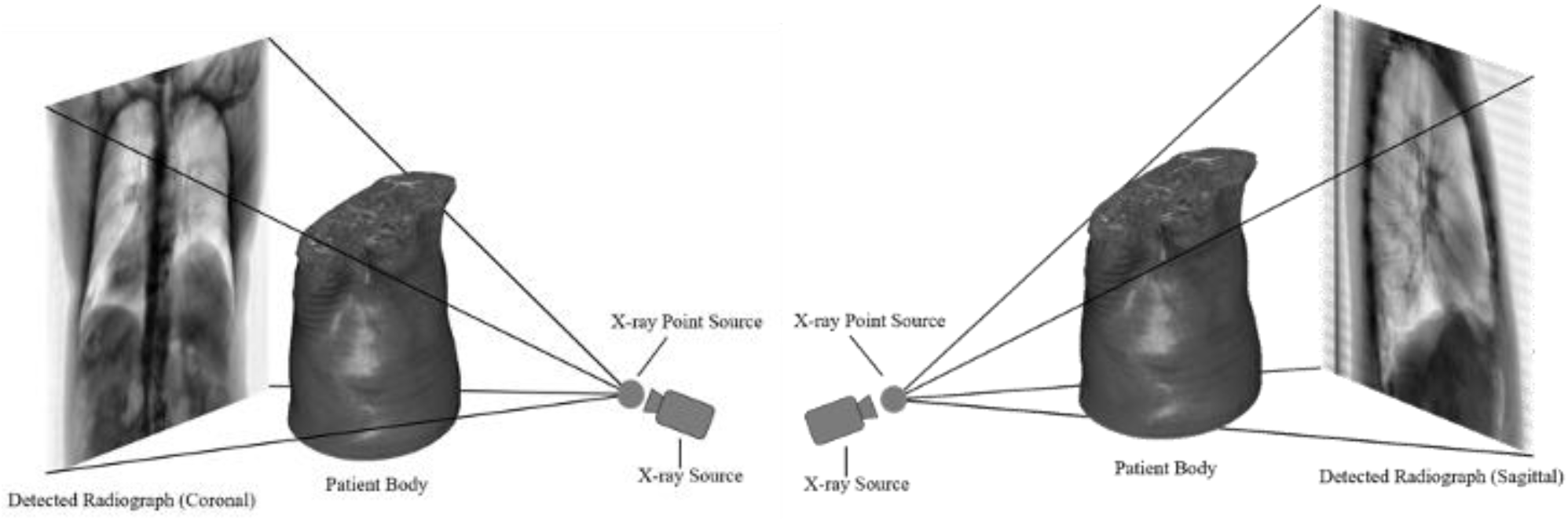
Illustration of the synthetic topogram creation process we applied to generate coronal (left) and sagittal (right) topograms from 1050 publicly available clinical CT volumes. The process involves simulating multiple x-ray beams that traverse through CT volumes from a fixed x-ray point source by utilizing the Beer-Lambert Law of attenuation, where final intensities are measured on a simulated detector with 1024 × 1024 pixels behind the patient body.

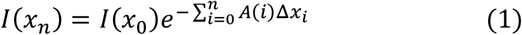

### B. Encoder-Decoder Neural Networks

Our formulated solution to the 2D-to-3D mapping problem is defined through a modified encoder-decoder neural network architecture with an embedded transformation subnetwork that is based on the autoencoder architecture^26^ (Figure 2). Consider an autoencoder *A* with input *X* and output *Y* such that *A*(*X*) = *Y*. Through stochastic backpropagation and a predefined loss function *L*(*X, Y*), the autoencoder iteratively updates the parameters of the neural network so that, given a representative input from the input dataset, the neural network may produce a near-identical output. We developed two architectures of encoder-decoder models which follow a similar methodology, one for a single-view input and one for a dual-view (stereo) input. We consider the problem of volumetric CT mapping from single or double 2D projection as a traditional image translation task with an additional transformation layer to increase image dimensionality. Given a coronal and/or sagittal topogram projections *X*_1_ and *X*_2_, the goal of the neural network is to generate a predicted output volume *Y*_*pred*_ such that *Y*_*pred*_ = *Y*_*truth*_, where *Y*_*truth*_ is the ground-truth clinical CT volume. We therefore define two deep learning mapping functions *F*_1_ and *F*_2_, such that *F*_1_(*X*_1_) = *Y*_*pred*_ and *F*_2_(*X*_1_, *X*_2_) = *Y*_*pred*_. Via stochastic gradient descent and convolutional backpropagation, we iteratively update the model weights of *F*_1_ and *F*_2_ according to the ground-truth CT volume *Y*_*truth*_ through a loss function *L*(*Y*_*pred*_, *Y*_*truth*_).

### C. Representation Subnetwork

The representation subnetwork (shown with yellow background in Figures 1 and 3) is tasked with reducing the dimensionality of the input 2D topogram or topograms. Given a single-view input topogram *X*_1_, or two dual-view topograms *X*_1_ and *X*_2_, a representation subnetwork is trained to generate a single output latent tensor *L* for each of the input topograms. The data flow of the subnetwork in both the single-view and dual-view neural networks is given as: 1024 × 1024 × 1 → 1024 × 1024 × 32 → 512 × 512 × 64 → 256 × 256 × 128 → 128 × 128 × 256 → 64 × 64 × 512 → 32 × 32 × 1024 → 8 × 8 × 4096 → 4 × 4 × 4096 with each ‘→’ representing a convolutional block with batch normalization and the Rectified Linear Unit (ReLU) activation function^27^. We chose the ReLU activation function over alternatives due to its faster convergence, and its tendency to resolve issues such as vanishing or exploding gradient, as found during initial tests.

**Figure 3:**
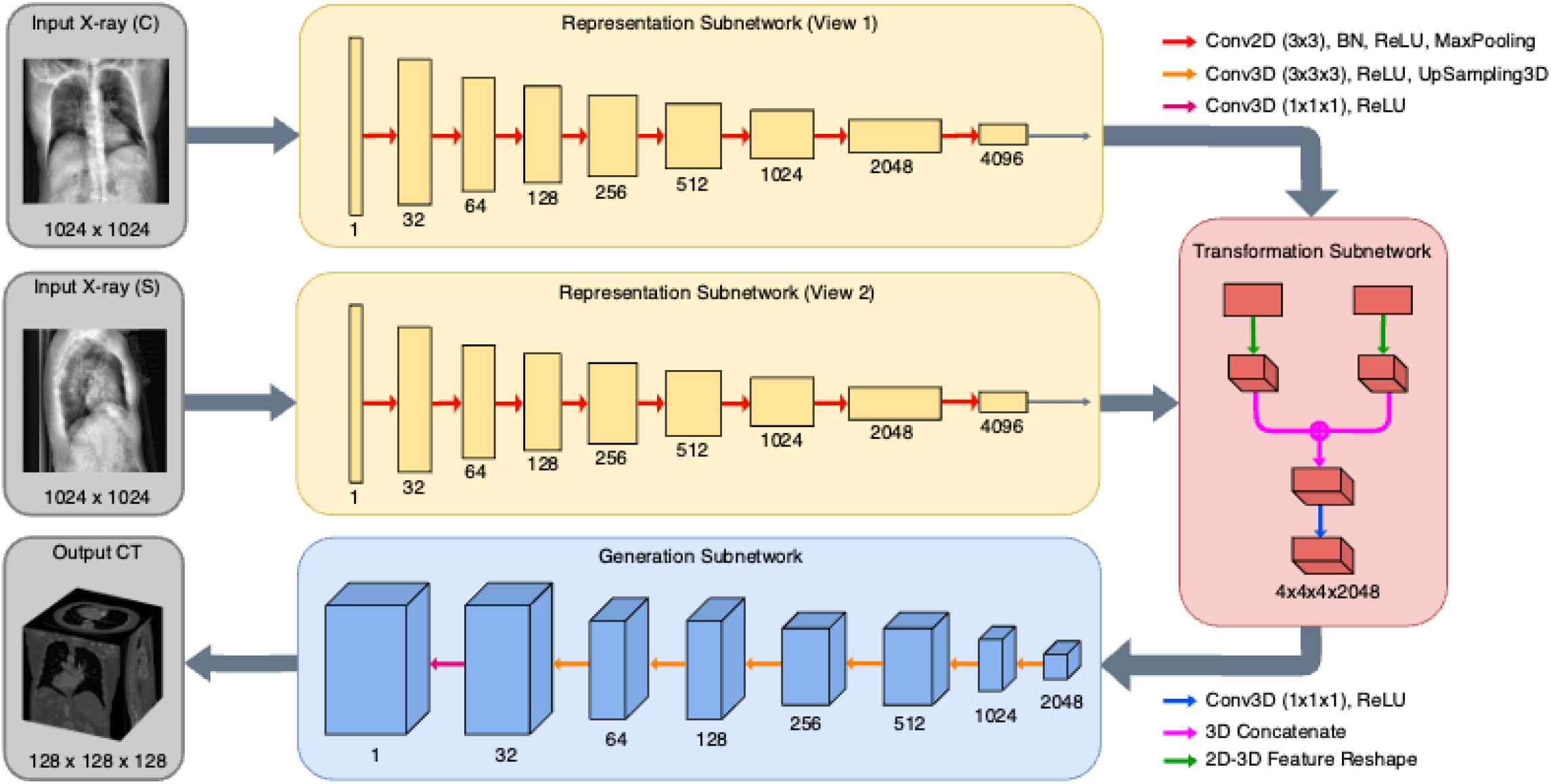
Architecture of a dual-input neural network, featuring four distinct modules: two representation subnetworks (yellow), a transformation subnetwork (red), and a generation subnetwork (blue). The number of channels is indicated by the number below each hidden layer. A coronal input view topogram (C) and a sagittal input view topogram (S), both with dimensions 1024 × 1024 × 1, are passed through separate but identical representation subnetworks and transformed into a volumetric output with dimensions 128 × 128 × 128. A similar architecture, with only a single representation network, is utilized for the dual-input neural network.

### D. Transformation Subnetwork

The transformation subnetwork (shown with red background in Figures 1 and 3) is tasked with combining the latent tensor representations of each radiographic projection and increasing their dimensionality. Considering transformation subnetwork *T*, latent tensor *L*, and reshaped latent tensor *Z*, the single-view subnetwork is invoked such that *T*(*L*) = *Z*. In the dual-view architecture, both previously generated latent tensors *L*_1_ and *L*_2_ are concatenated to form a higher-dimensionality latent tensor *Z*. Hence, we denote the operation as *T*(*L*1, *L*2) = *Z*. In both variants of the transformation subnetwork, a single convolution with kernel size 1 × 1 × 1 is invoked to learn the new reshaped spatial hierarchies.

### E. Generation Subnetwork

The generation subnetwork is the final component of the developed set of encoder-decoder neural networks and is tasked with enlarging the reshaped latent tensor *Z* into a final volumetric output. Considering the reshaped latent tensor *Z* and final output CT volume *Y*_*pred*_, the generation subnetwork, abstracted as *G*, is invoked such that *G*(*Z*) = *Y*_*pred*_. The data flow of the hidden convolutional layers in the generation subnetwork is given as: 4 × 4 × 4 × 1024 → 8 × 8 × 8 × 512 → 16 × 16 × 16 × 256 → 32 × 32 × 32 × 128 → 64 × 64 × 64 × 64 → 128 × 128 × 512 → 128 × 128 × 128 × 1, with each ‘→’ representing a deconvolutional block with the ReLU activation function.

### F. Network Training

To determine the accuracy of both single-view and dual-view architectures, both neural networks are trained on the paired synthesized topogram and volumetric CT dataset. For both models, an initial learning rate of 0.0002 on the Adam optimizer is used to minimize the mean-squared-error (MSE) loss function *L*(*Y*_*pred*_, *Y*_*truth*_) via stochastic gradient descent and backpropagation. All training was conducted on two SLI-connected NVIDIA Tesla P100 GPUs, each with 16 GB of VRAM. Model weights are saved locally every 10 epochs, and the training process automatically terminates after convergence. The weights with the lowest average loss are preserved and serialized.

### G. Evaluation Metrics

For quantitative evaluation, four common image similarity metrics are calculated: mean-squared error (MSE), mean average error (MAE), structural similarity (SSIM), peak signal-to-noise ratio (PSNR). Additionally, accuracy of lung/tissue segmentations that are based on thresholding the volumes produced by both neural networks and the original (ground-truth) CT volume is used as an application-focused metric with DICE score calculations^28,29^. Finally, we adopt a methodology to determine the quality of the recovered volumes as input for dose modulation techniques. For this, we first remove extraneous objects from the CT volume through thresholding and connected component labeling^30^. Next, we calculate the water equivalent diameter (*D*_*w*_) of each slice on the ground-truth and estimate volume slices by:

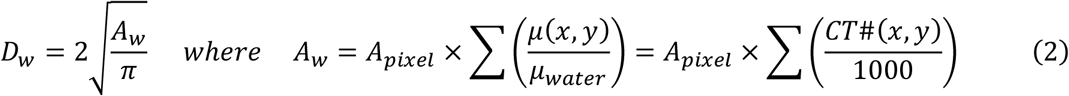

where *CT*#(*x, y*) represents the water-normalized attenuation of a voxel situated at coordinates (*x, y*), *A*_*pixel*_ represents the area of a pixel, and *A*_*w*_ represents the water-equivalent area of the given CT slice. This method with its underlying approximations was demonstrated to be valid in Wang *et al*. with both analytical and Monte Carlo methods^31^. Similar calculations and are also included as part of the AAPM task group 220 Report^14^. In addition to these quantitative image similarity metrics, qualitative analysis was performed on typical generated samples.

## III. Results

The results reported in the section were averaged over all 60 test dataset volumes, i.e., CT cases that were never seen by either of the neural networks, to determine the efficacy of the proposed deep learning framework for dose modulation and patient size estimation tasks.

Figures 4 and 5 display example results of the two neural networks as compared to their corresponding ground-truth CT datasets. Error maps of intensity difference between original CT volumes, representing ground-truth intensities, and 3D volumes based on 2D inputs demonstrate considerably lower errors for volumes generated from dual-view inputs compared to those generated from single-view inputs. These observations are consistent with the lower MAE and MSE values of the dual-view neural network, as given in Table 1. In addition, a significantly lower error is observed at the patient boundary in both error maps and true-positive/false-positive/false-negative maps. These results indicate that the dual-view neural network is more adept than the single-view neural network at recovering the boundaries of the thoracic anatomy of the patients, an important consideration for advanced scan planning applications. Specific examples of areas where the dual-view neural network outperforms the single-view neural network can be observed in both the left and right upper apicoposterior lobes, as well as the medial and anterior lower lobes. Additionally, the trachea and superior mediastinum regions shown are almost fully recovered by both models. The dual-view neural network recovered the trachea almost perfectly, while the single-view neural network leaves several boundary voxels omitted. While soft tissue anatomies are recovered accurately, we note a generally poor recovery of contrast and details of boney tissue that is most evident in the spine. That said, the dual-view neural network does demonstrate more promise in recovering both soft and boney tissues.

**Table 1:** Image similarity metrics for single- and dual-view neural network architectures. While both networks demonstrate low errors and high correspondence with the ground-truth CT datasets, there is an apparent advantage for the dual-view network for all matrices.

| Architecture | MAE | MSE | SSIM | PSNR | DICE |
| --- | --- | --- | --- | --- | --- |
| Single-view | 0.0013 | 0.0012 | 0.8893 | 29.4531 | 0.9401 |
| Dual-view | 0.0004 | 0.0009 | 0.9396 | 31.5240 | 0.9676 |

**Figure 4:**
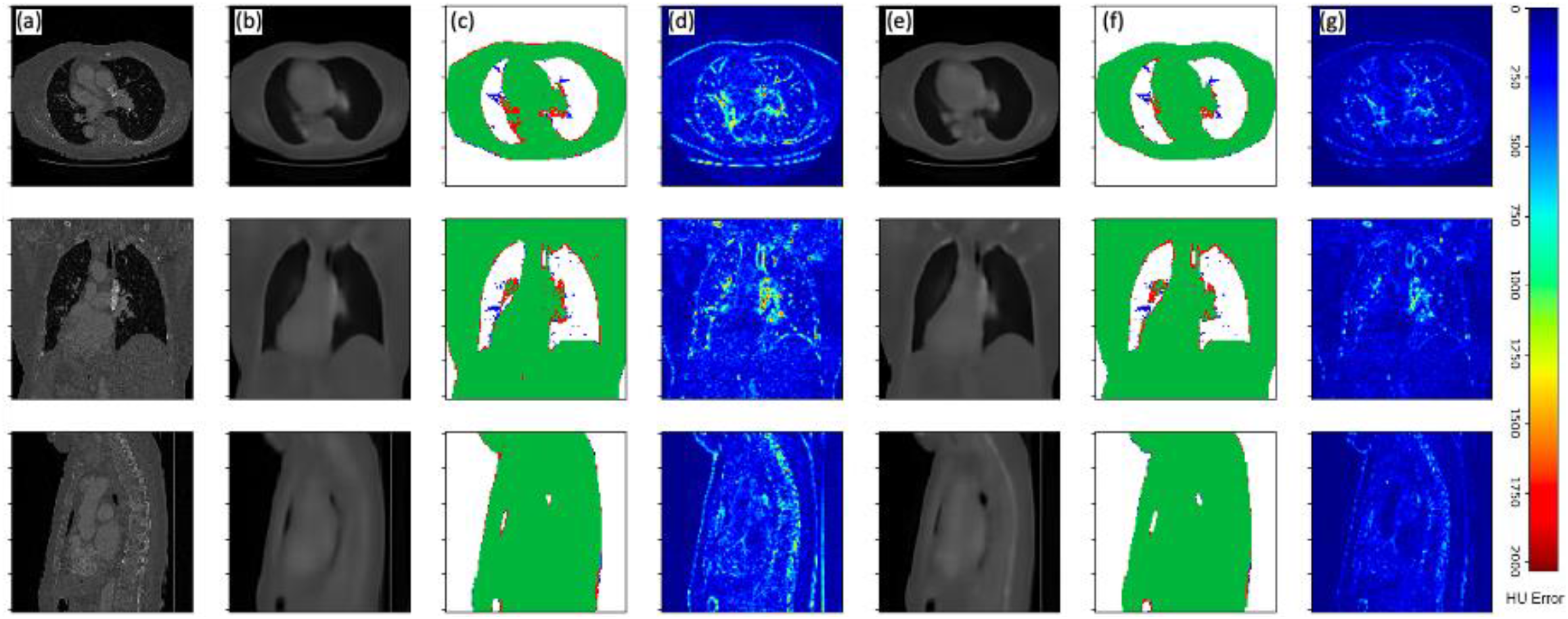
Volumetric surview (3D-surview) results for an example CT dataset of a single patient viewed in axial (top row), coronal (middle row) and sagittal (bottom row) anatomical planes. Displayed columns are (a) ground-truth CT slices, (b) slices generated by the single-view neural network, (c) a true positive (green), false positive (blue), and false negative (red) map for single-view volume generation, (d) absolute HU error map for single-view volume generation, (e) slices generated by the single-view neural network, (f) a true positive (green), false positive (blue), and false negative (red) map for single-view volume generation, (g) absolute HU error map for single-view volume generation.

**Figure 5:**
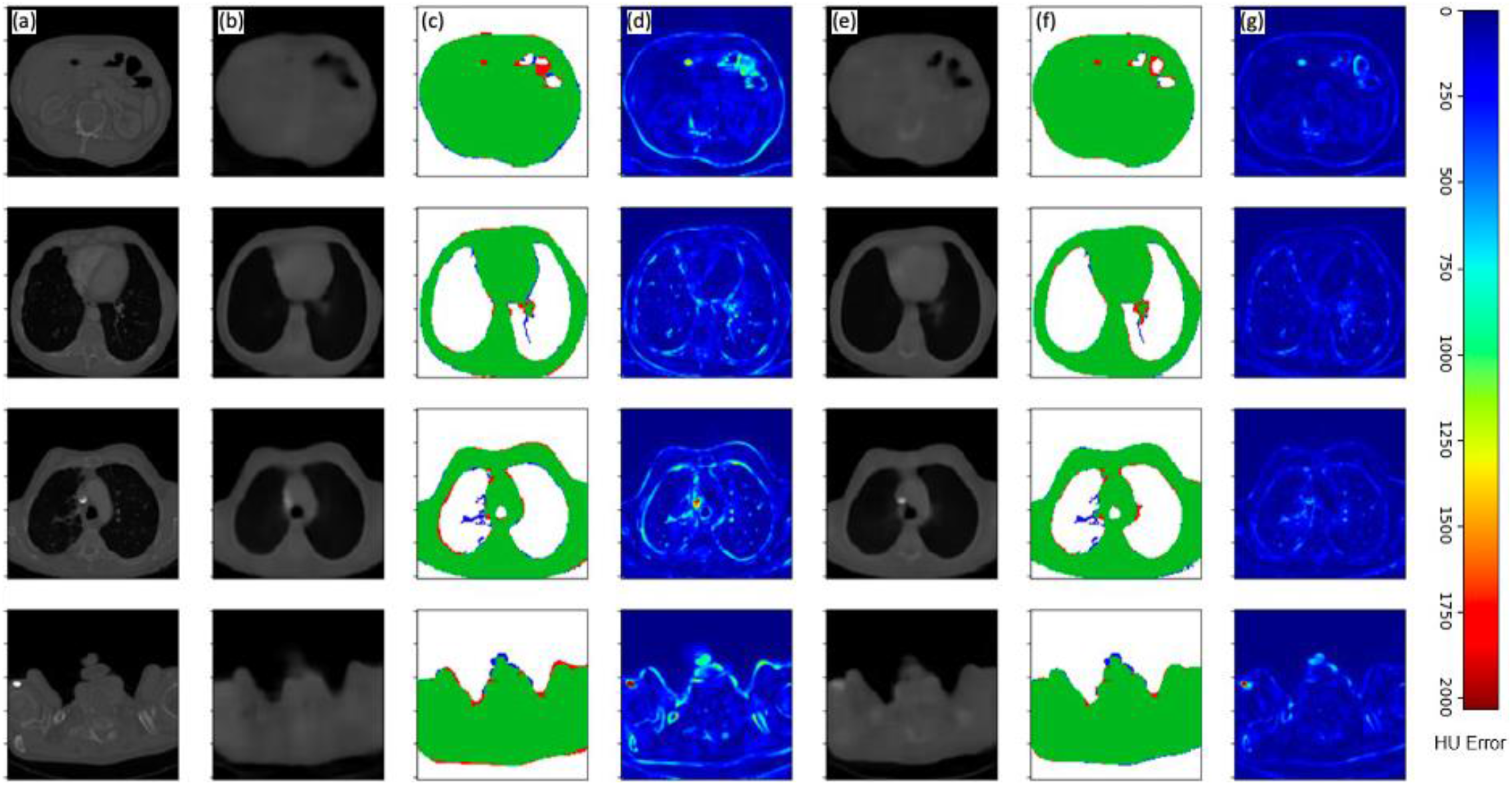
Volumetric surview (3D-surview) results for an example CT dataset of a single patient viewed in multiple axial slice depths (different rows). Displayed columns are (a) ground-truth CT slices, (b) slices generated by the single-view neural network, (c) a true positive (green), false positive (blue), and false negative (red) map for single-view volume generation, (d) absolute HU error map for single-view volume generation, (e) slices generated by the single-view neural network, (f) a true positive (green), false positive (blue), and false negative (red) map for single-view volume generation, (g) absolute HU error map for single-view volume generation.

The lungs in a CT volume can be (naïvely) segmented by thresholding and applying connected component analysis ^30^. Comparisons of lung segmentations of neural network generated volumes and of lung segmentations from ground-truth CT volumes reveal that both networks result in accurately segmented lungs, with DICE scores of 0.9555 and 0.9789 for the single and dual-view neural networks, respectively (Table 1). The high-fidelity lung/tissue-segmentations can also be perceived from true-positive (TP) / false-positive (FP) / false-negative (FN) error maps for both models, where the volumes generated from dual-view neural network present fewer false-negative and false-positive voxels than the volumes generated from the single-view neural network.

Examples of patient sizes estimations, as represented by water equivalent areas (*A*_*w*_) distributions along the patient axis, are displayed in Figure 6. Note the similarities in mesh geometries between the ground truth representation (a), the single-view neural network representation (b), and the dual-view neural network representation (c). Calculated average water equivalent diameters (*D*_*w*_) resulted in accurate patient size estimations for both neural networks, with the single-view architecture scoring 0.911 and the dual-view

**Figure 6:**
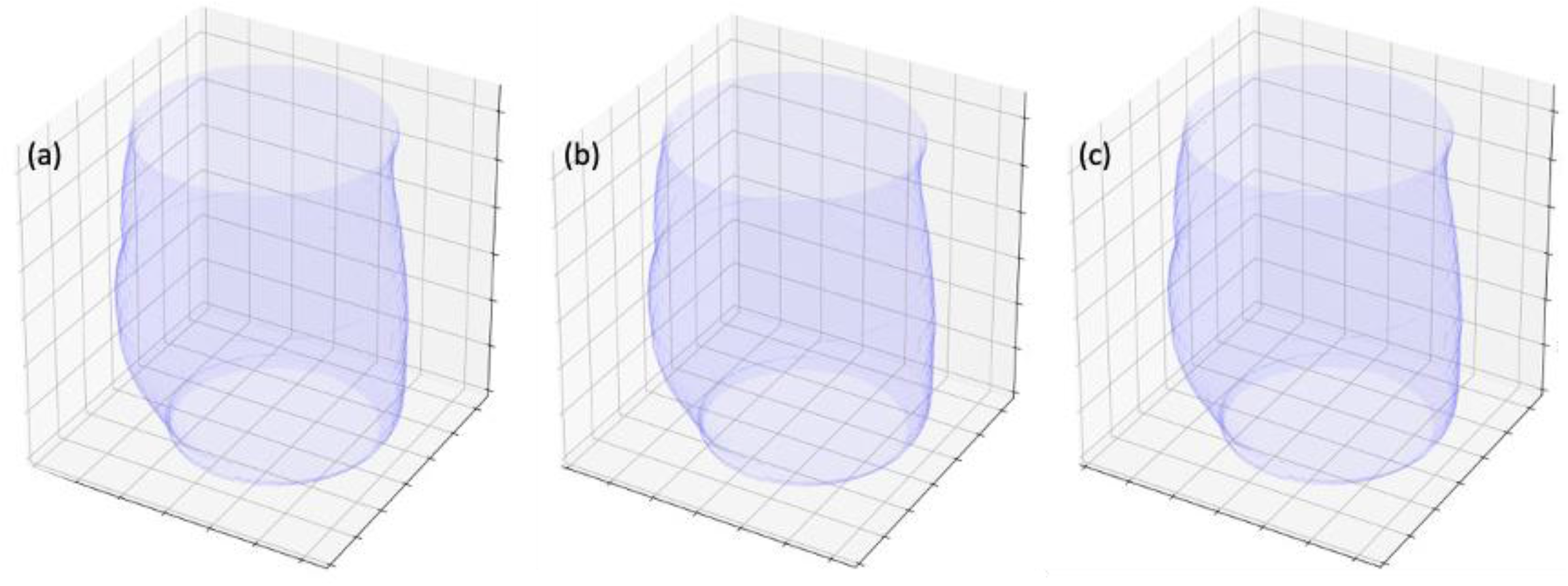
An example of volumetric external bodies and volumetric water equivalent areas (*A*_*w*_) for ground-truth datasets and neural network generated 3D-surview volumes. Displayed are (a) meshes created from the water-equivalent-areas of a ground-truth CT volume, (b) a CT-like 3D-surview generated by the single-view neural network, and (c) a CT-like 3D-surview generated by the dual-view neural network. Our results demonstrate excellent recovery of body size representations for a variety of applications, e.g., radiation dose modulation.

architecture scoring 0.925. Based on these results, the two neural networks, and especially the dual-view variant, demonstrate a potential to improve dose modulation techniques by providing accurate volumetric information as input for calculating efficient dose-modulated CT acquisitions.

## IV. Discussion

In this investigation, we develop and refine two encoder-decoder neural network architectures for recovering volumetric CT data from single- or dual-view radiographic projections. The developed models were trained and tested on paired data of publicly available CT datasets and synthetically generated CT tomograms (Figure 2). While the generated 3D-surviews do not contain sufficient information for diagnostic tasks and are thus not considered as an alternative to volumetric CT datasets, both the single and the dual-view networks perform well on image similarity and use-case specific metrics. Moreover, our results indicate that, in most cases, the additional information that is available with the dual-view neural network enables improved accuracy as compared to the single-view neural network. Specifically, from qualitative observations and quantitative image similarity metrics across multiple samples, we acknowledge that the dual-view neural network is generally superior at recovering both the anatomy and soft-tissue/bone intensities than the single-view neural network and is thus more relevant for further clinical translation developments. However, both models perform comparably to the current state-of-the-art in deep learning CT recovery from limited information ^21–23^, indicating the viability of both architectures in practice.

Based on our qualitative evaluation, the DICE scores from Table 1, and the water-equivalent area and diameter comparisons (Figure 6), we deduce that there are only minor differences between patient size estimations that rely on the complete information available with the (retrospective) diagnostic scan and patient size estimations that rely on information that was extracted by the single or dual-view neural networks. Our results indicate that the developed deep neural networks can provide a promising method with potential applications for automatic acquisition parameter selection^32^, e.g., automatic kVp selection ^12,13^, more efficient dose modulation techniques that utilize improved high-dimensional input, e.g., for various dynamic bowtie filter concepts^15–17^, and improved scan planning with better anatomy-awareness. Tube current modulation during diagnostic CT scans is crucial a technique that allows a significant reduction of ionizing radiation dose, and thus improved patient safety, while maintaining diagnostic image quality. Conventionally, metrics such as patient size, measured as water-equivalent diameter, or local (along the patient axis) cylindrical or elliptical cross-section representations, also measured in water-equivalent diameter, are used to calculate position/angle-dependent radiation dosages^14^. In addition, calculated patient sizes provide the basis for size-specific dose estimates^14^ (SSDE). Finally, the additional information may be of sufficient resolution to enable sufficiently accurate tissue attenuation inputs for the generation of PET attenuation correction maps^33^ with a significant reduction of ionizing radiation dose levels, i.e., use of single- or dual-view tomograms instead of a complete volumetric scan.

In conclusion, we demonstrate an ability to utilize low-dose single- or dual-view CT topograms, which in the current clinical practice are acquired in almost all CT examinations, for estimating volumetric patient anatomies with high accuracy. Due to the abundance of matched CT-topogram data, such neural networks can be trained and integrated into all existing CT scanners relatively seemingly, i.e., without requiring any hardware changes, to enable acquisition parameter recommendations or automatic selections, a reduction in ionizing radiation dose levels through improved dose modulation calculations, and have a potential to enable PET attenuation correction maps with only a fraction of the currently utilized radiation dose.

## Data Availability

All data produced in the present study are available upon reasonable request to the authors

## V. Acknowledgment

We acknowledge support through the National Institutes of Health (R01EB030494) and Philips Healthcare.

## Notes

### Author Declarations

Only existing public datasets were used

